# Paediatric Dentistry and the coronavirus (COVID-19) response in the North East of England and North Cumbria

**DOI:** 10.1101/2020.06.02.20114967

**Authors:** S Simpson, O Sumner, R Holliday, CC Currie, V Hind, N Lush, LA Burbridge, BOI Cole

**Affiliations:** School of Dental Sciences, Faculty of Medical Sciences, Newcastle University, Newcastle upon Tyne; Newcastle upon Tyne Hospital NHS Foundation Trust, Newcastle upon Tyne

## Abstract

**Introduction:** Coronavirus (COVID-19) has dramatically changed the landscape of dentistry including Paediatric Dentistry. This paper explores paediatric patient data within a wider service evaluation completed within an Urgent Dental Care Centre in the North East of England and North Cumbria over a 6-week period.

**Aim:** To assess demand for the service, patient demographics and inform paediatric urgent dental care pathways.

**Main outcome methods:** Data collected included key characteristics of paediatric patients accessing Paediatric Dental Services from 23^rd^ March to 3^rd^ May 2020. Descriptive statistics were used for analysis.

**Results:** There were 369 consultations (207 telephone, 124 face-to-face and 38 Out of Hours consultations). The mean age of children accessing the service was 7 years old. 7% of those attending face-to-face visits were reattenders. The most common diagnoses were irreversible pulpitis and dental trauma. 49% of face-to-face consultations resulted in extractions, 28% with General Anaesthetic, and 21% with Local Anaesthetic.

**Conclusion:** Management of dental emergencies provided by the Urgent Dental Care Centre for paediatric patients has largely been effective and confirmed the efficacy of patient pathways established.

Three in Brief Points
⍰ Describes the approach adopted in the North East of England and North Cumbria to managing paediatric dental emergencies during the coronavirus pandemic
⍰ Provides an overview of dental problems and management provided to paediatric patients in the first 6 weeks of the coronavirus pandemic
⍰ Confirms the need for general anaesthetic services for exodontia in the paediatric population

## Introduction

On the 25^th^ of March 2020, the Chief Dental Officer of England directed all General Dental Practices to cease all patient facing activity to help control the spread of the novel coronavirus SARS-CoV-2.^1^ This triggered the radical restructuring of dental services and the establishment of Urgent Dental Care Centres (UDCC).

The primary source of transmission of COVID-19 appears to be respiratory droplets.^2^ The virus has also been shown to survive on hard surfaces for a significant time period.^3^ It is thought that the virus can be transmitted via contact with asymptomatic individuals.^4^ Furthermore, within dentistry transmission can occur through inhalation from aerosol generating exposures such as coughing or sneezing and aerosol generating procedures (AGPs).^5^

In England 23.3% of 5-year-old children have experience of dental decay, with on average of 0.8 teeth affected per child.^6^ The North East of England is similar to the national average with 24% of children having obvious dental decay.^6^ In addition, approximately 12% of 12-year-olds and 10% of 15-year-olds have evidence of dental trauma to permanent incisors.^7, 8^ This indicates a significant level of dental disease and treatment need in the paediatric population in the North East of England. It can therefore be assumed that there will be an ongoing need for urgent dental care for this population despite the ongoing pandemic.

During the first 6 weeks of the UK Coronavirus pandemic, the Paediatric Dental Service, based in the Dental Hospital of Newcastle upon Tyne NHS Hospitals Foundation Trust (NuTH), operated a specialist led 7-day emergency dental service with weekly access to General Anaesthesia (GA) exodontia lists to manage dental emergencies in the paediatric population. The aim of the service was to minimise risk of repeat attendance and maximise treatment efficiency. To the authors’ knowledge this was the only 7-day specialist led service of its kind operating in England.

Historically the specialist Paediatric Dental Service at Newcastle Dental Hospital had operated a walk-in urgent care service for children ages 0-16 years. This service was suspended immediately due to the Government’s social distancing guidelines, as it could not safely operate in its traditional structure. An email urgent care referral system was set up alongside the adult services in the region which accepted referrals from General Dental Practitioners, NHS 111, Community Dental Services and Hospital Medical Services, and, self-referrals for a limited range of dental problems as detailed in Figure 1.

**Figure 1.**
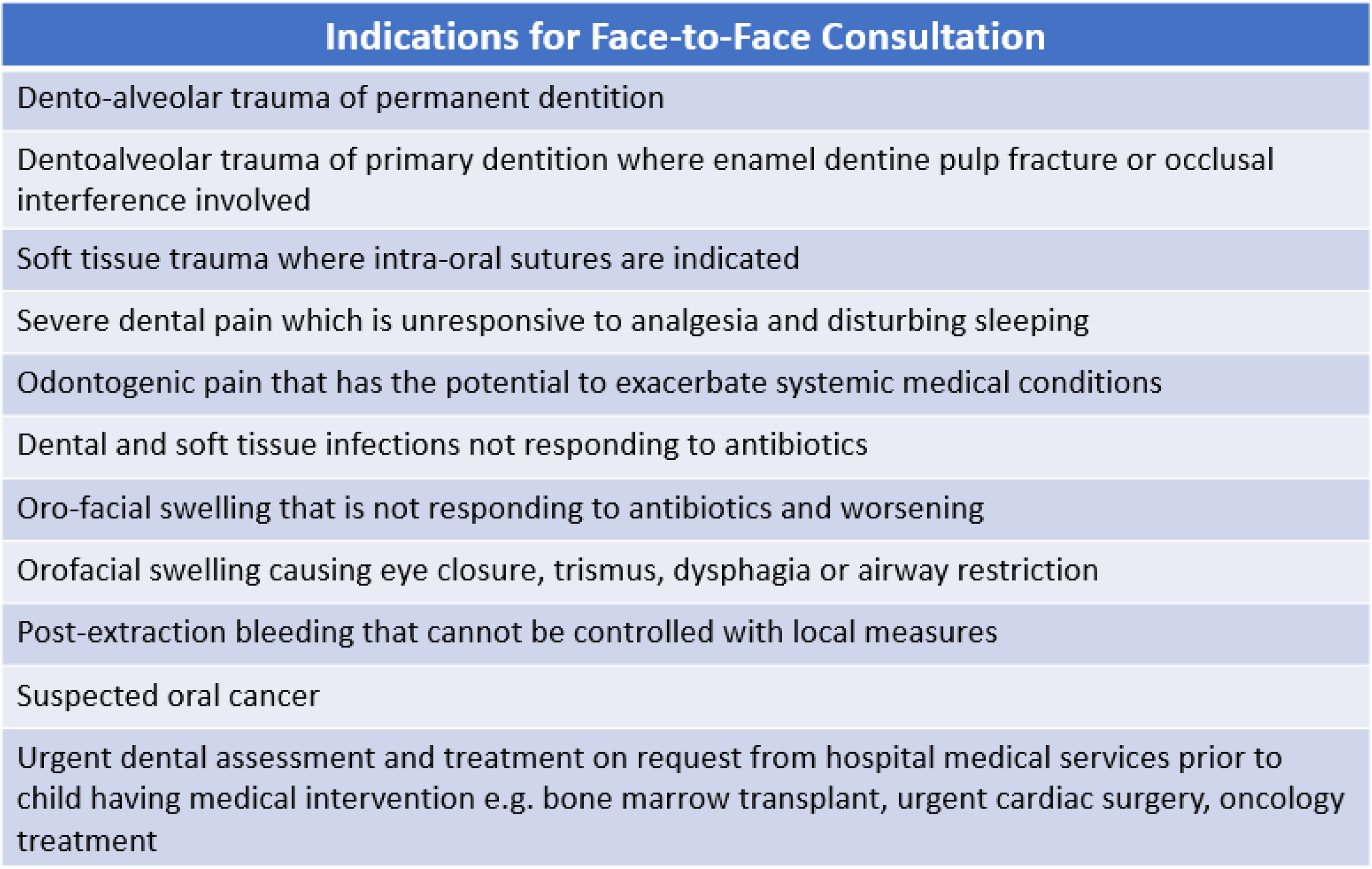
Description: Dental problems accepted for face-to-face consultation

This service operated in line with all aspects of the Royal College of Surgeons of England National Guidance for Paediatric Dentistry during the COVID-19 pandemic despite our UDCC being established before the guidance was published.^9^ The Newcastle Dental Hospital UDCC was the sole provider of face-to-face urgent dental care in the region, during the first 4 weeks following mass closure of General Dental Practices. A service evaluation, covering adult and paediatric services, has already been published.^10^ Paediatric dentistry demands, outcomes and service considerations are worthy of further consideration and were not presented in the previous publication. This paper aims to explore and discusses these in greater detail, considering the main challenges and key learning points to inform future practise.

## Methods

The data presented in this paper represents the paediatric patient data of a previously published service evaluation.^10^ The methods have previously been described in detail but in summary involved data collection by clinicians as part of a service evaluation of the Newcastle Dental Hospital UDCC during the COVID-19 pandemic (NUTH Clinical Effectiveness Register reference 1006). Data included basic demographics (gender, age, partial postcode, COVID-19 status), patient pathway details (referral source, repeat attendance) and outcomes. Descriptive statistics were performed in SPSS (Windows version 25.0.0.1; SPSS Inc., Chicago [IL], US).

## Results

The Paediatric Dental Service provided 369 consultations, of which 56% were telephone triage consultations (n=207), 34% (n=124) were face-to-face consultations as part of the in-hours service and 10% (n=38) as part of the out-of-hours service. The mean age of children receiving a telephone triage consultation was 7.0 years old (range 0-16 years, SD 3.4 years); 48% were female (n=100) and 52% male (n=107). The majority of patients who accessed the triage service reported having a GDP (n=179, 86%), and out of these, 125 (70%) had attempted to contact them before contacting the UDCC; 17 (9%) had received no or inadequate advice. There were a small number (n<5) of alternative referral routes such as referred for urgent dental assessment and treatment by medical specialties at the Great North Children’s Hospital (NuTH). The most common reasons for referral were irreversible pulpitis (33%, n=69) and dental trauma (24%, n=50). Less common reasons for referral, grouped together as ‘other’, included conditions specific to Paediatric Dentistry such as neonatal teeth (n=2) and unusual occurrences such as foreign object stuck between teeth/in gingivae (n=2). Figure 2 illustrates the telephone triage diagnosis.

**Figure 2.**
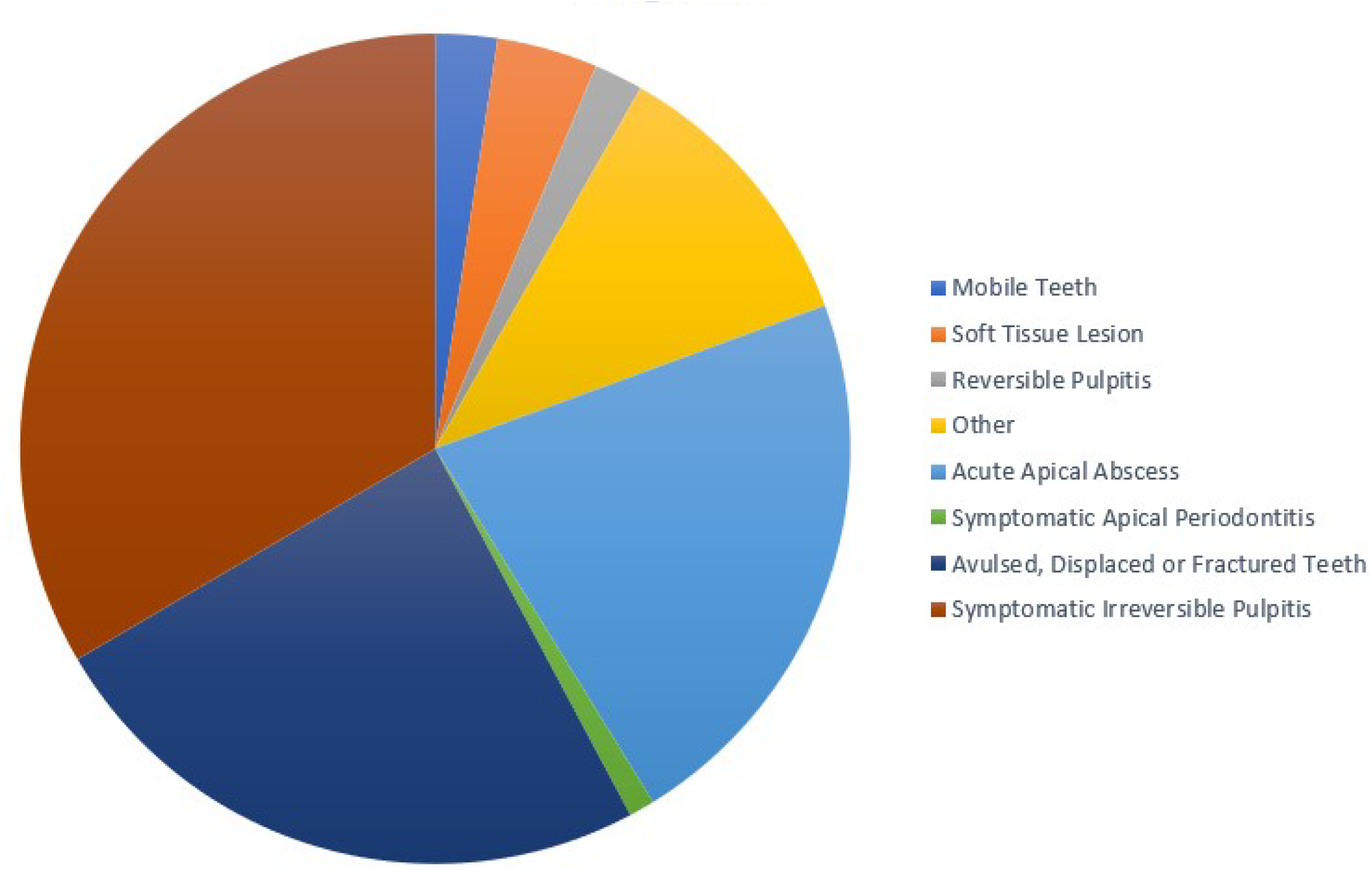
Description: Frequency of dental diagnosis of patients referred to UDCC for telephone and face-to-face consultations at the Newcastle Dental Hospital during the COVID-19 pandemic. ‘Other’ notable diagnoses included: Patient queries (n=13); lost preformed metal crown (n=1); urgent dental assessments as requested by medical specialties (n=3); non-odontogenic extra-oral swelling (n=1); foreign object stuck between teeth/in gingivae (n=2); neonatal teeth (n=2); traumatic occlusion (n=l).

The vast majority of those accessing the triage service had no symptoms of COVID-19 (71%, n=148). Eight children were symptomatic for COVID-19, with very few of these requiring face-to-face consultations (the exact figure cannot be reported in the interest of protecting patient confidentiality). Eight percent of children (n=13) attending face-to-face consultations were in a shielding category.^11^

The majority of patients that received a face-to-face consultation were first time attenders (76%, n=123) with 7% (n=11) attending for a repeat visit. The reasons for repeat attendance included: parent declined treatment (n=1); incomplete extraction (n=2); assessment and planned return for extractions under GA (n=6); debonding of reattached tooth fragments/adhesive bandages (n=2). Patients travelled a mean distance of 29km (range 3-142km, SD 25km). This peaked in week 3 at 39 km and reduced to 25-26 km in weeks 5-6 (when other UDCCs were established). The mean age of patients attending face-to-face consultation were comparable to those that received telephone triage consultations (7.0 years old, range 0-16 years, SD 4.0 years).

Half of the face-to-face consultations resulted in extractions (49%, n=80) with either local anaesthetic (21%, n=35) or general anaesthetic (28%, n=45). Overall, 36% of patients (n=59) were successfully managed with local anaesthesia alone (extractions, management of trauma in the permanent dentition and extirpations). All the patients (n=22) referred with trauma in the permanent dentition received a face-to-face assessment. Less than 5 AGPs were deemed necessary in the first 6 weeks of this service, the exact figure was very low and therefore cannot be reported to protect patient confidentiality. Figure 3 demonstrates the outcomes of patients attending face-to-face consultations.

**Figure 3.**
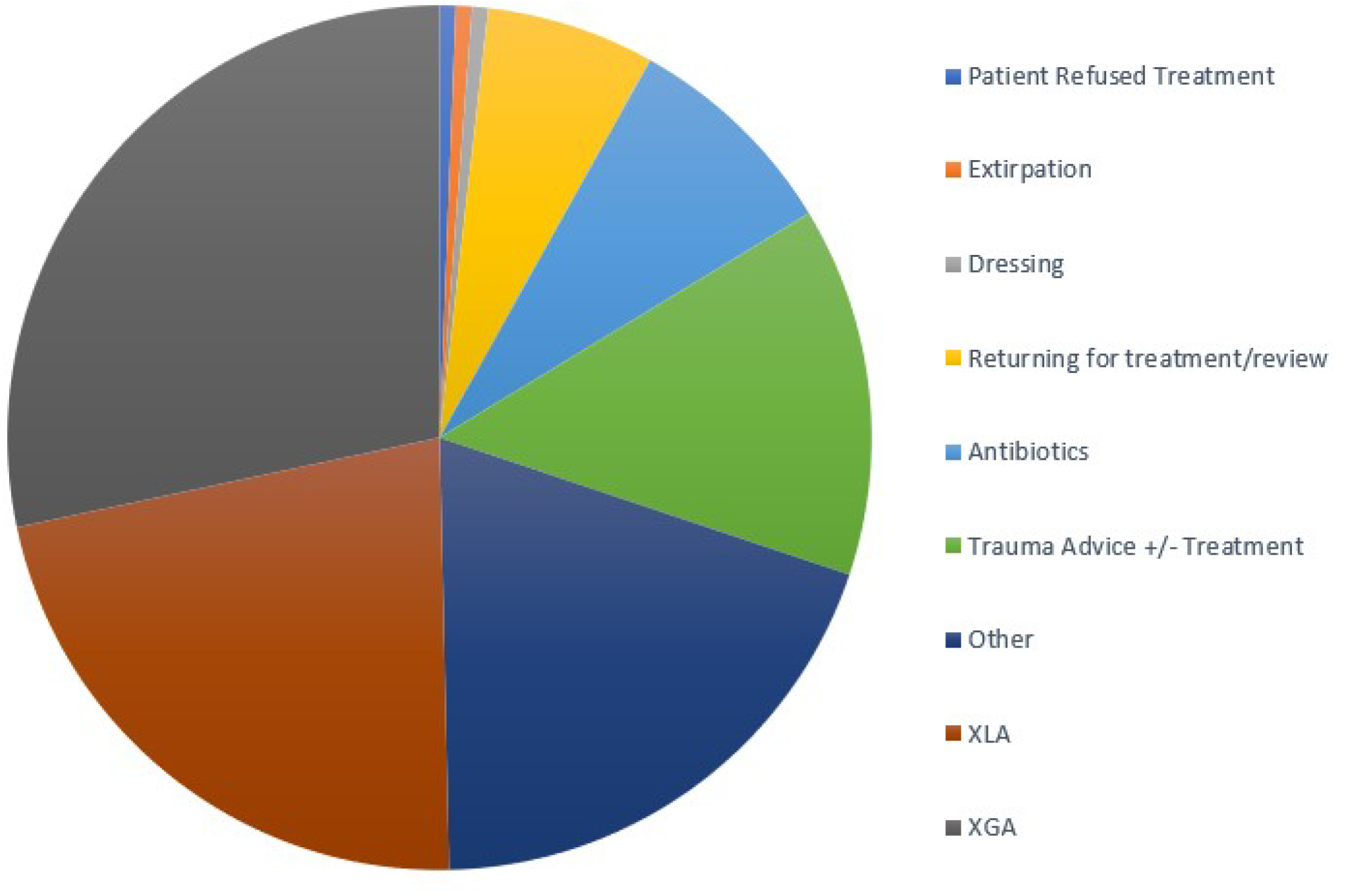
Description: Frequency of face-to-face consultation outcomes of patients attending the Newcastle Dental Hospital UDCC for treatment. ‘Other’ notable outcomes include: pre-oncology assessment (n=2); Pre-cardiac assessment (n=1); removal of foreign object from teeth/gums (n=2).

## Discussion

This service evaluation provided a review of paediatric urgent dental care patient pathways to inform quality improvement and workforce planning. It has provided a unique opportunity to assess urgent dental care needs of the local population and outcomes of a 7-day specialist led service. The outcomes to date indicate the protocols for management of acute dental problems within the paediatric population during the COVID-19 pandemic have been largely effective with few repeat attendances. It has also given us the opportunity to document the distance paediatric patients require to travel within the region for specialist care which will be useful for informing specialist service provision after the pandemic has resolved.

Very few patients with COVID-19 symptoms had a clinical need that justified face-face-attendance with the majority of patients referred to the service asymptomatic.^12^ A small proportion were shielding due to medical histories increasing patient vulnerability to COVID-19.^11^ AGPs were largely avoided however when required were carried out in a designated AGP closed surgery with full Personal Protective Equipment (PPE) comprising of theatre cap, long sleeved surgical gown, FFP3 mask, visor, and gloves. ^13, 14^

A specific consideration and challenge for paediatric dentistry has been the need to manage dental trauma of the permanent dentition whilst avoiding AGPs. In developing permanent incisors, loss of vitality has a profound impact on complexity of treatment need and overall prognosis. Furthermore, co-operation for treatment in the age range of the developing permanent incisor can be very variable adding to the overall complexity of treatment. All dental traumas of the permanent dentition referred to the service received face-to-face consultation and management to maximise treatment outcomes.

The incidence of trauma to permanent incisors in the evaluation sample was greater than the reported national average of 1 in 10 children experiencing dental trauma to permanent incisors.^8^ Lack of school attendance may account for this where children would ordinarily spend 6-7 hours per weekday carrying out indoor academic, low energy activities. Anecdotally, trauma in the evaluation sample had been caused with outdoor, high-energy activities such as accidents on trampolines, scooters, and bicycles.

The current SDCEP guidance on Management of Acute Dental Problems During COVID-19 pandemic, does not classify enamel dentine fracture in permanent incisors as an indication for face-to-face management. ^12^ Literature suggests that pulpal necrosis can occur in up to 40% of enamel dentine fractures without dentine protection.^15^ In the event of pulp necrosis, in the current pandemic there is the potential for delayed extirpation with inflammatory resorption as a potential sequela. In immature permanent incisors this can be devastating and render a tooth unrestorable. Dentine protection can be achieved with self-etching bond and flowable composite or even glass ionomer cement avoiding the need for an AGP. It would therefore seem reasonable to seal exposed dentine in enamel dentine fractures of the immature incisor in asymptomatic children during the COVID-19 pandemic.

Delayed repositioning of displacement injuries can result in difficulty with surgical repositioning. Furthermore, there is an association between poorer outcomes and delayed treatment with literature recommending that luxation and displacement injuries are managed within 24 hours.^16^ Specialist/consultant level clinicians were on a weekend rota to allow prompt management of complex dentoalveolar trauma.

Dental anxiety in this patient population is an ingrained challenge for the specialty. Anecdotally, we found this to be heightened in both parents and patients due to added fear of catching coronavirus. Furthermore, we are unable to provide inhalation sedation to date due to difficulties disinfecting the reusable nasal hoods and tubing. Behavioural management techniques were however effectively employed in over a third of patients attending face-to-face assessment to successfully manage extraction and trauma management under local anaesthetic.

We were extremely fortunate to have availability of GA throughout the coronavirus pandemic to manage children who could not tolerate treatment in the dental chair. A need for access to GA services in the region has been supported with 28% of face-to-face consultations resulting in extractions under GA. Since completion of the service evaluation, demand for GA has increased with 4 GA lists operating per week where availability allowed.

Changes to Standard Infection Control Procedures, particularly the move to use chlorine disinfectant to clean clinical areas has resulted in a reduction in the number of patients that can be seen in a clinical session.^13^ This had a significant impact on the GA service, increasing procedure time for exodontia under GA, effectively halving the number of patients that can be accommodated on a GA list.

A small number of children were referred by paediatric oncology and cardiology to the Paediatric Dental Service during the service evaluation. In our normal service, we have close links with medical and surgical colleagues in the Great North Children’s Hospital (Newcastle Upon Tyne NHS Trust) including cardiology, oncology, haematology and immunology. We are often asked to provide urgent dental assessments for children prior to surgery or commencement of radiotherapy, chemotherapy, immune modulating agents and Bone Marrow Transplantation. Given the significant risk to the child’s health from odontogenic infection, this service was prioritised and continued.

To ensure children were dentally fit for their medical management, we facilitated dental extractions under GA within the Great North Children’s Hospital or as a piggyback procedure on planned surgical procedures. Where dental extractions were not appropriate, the risk of odontogenic infection from proceeding with medical management with untreated dental disease was clearly communicated and discussed with medical colleagues.

The data and experiences presented in this paper represent the experiences of the Urgent Dental Care Centre in the Newcastle Dental Hospital (Newcastle upon Tyne NHS Hospital Foundation Trust).^10^ This was shaped by the patient demographics, geographical and cultural factors specific to our region as well as organisational structure of the Paediatric Dental Service and UDCC. This should be taken into account when generalising the findings to other settings and geographical locations. COVID-19 status of patients was based on presence or history of symptoms as a result it may not be a true representation of COVID-19 status in the patient population assessed and should be interpreted with caution and within this context. Our anonymised data collection meant that we were unable to state the exact number of individual patients seen within the service, and instead we report the number of consultation episodes.

The future is likely to involve specialist paediatric services operating at full capacity, families travelling significant distances to access specialist services with long intervals between appointments and associated financial and social implications for families. In fact, these have all been suggested as reasons to establish managed clinical networks (MCNs).^17^ As dental services restart and resume in the post-COVID-19 era, this could present a prime opportunity for commissioners of services to accelerate development of MCNs.

## Conclusion

This service evaluation has given an insight into the urgent dental care needs of the paediatric population in the North East of England and North Cumbria. Irreversible pulpitis and dental trauma were most common reason children accessed the service largely resulting in extractions. The need for maintaining GA services and services to support the oral health needs of children with complex medical needs was evident. The need for AGPs were largely avoided despite the demand to manage dental trauma. Most children presenting to the service had no symptoms of COVID-19. With the potential for similar scenarios in the future, this data can be used to support the restructuring of Paediatric Dental Services and improve preparedness to meet future challenges.

## Data Availability

No external dataset.

## Acknowledgements

The authors declare no potential conflict of interest with respect to authorship and/or publication of this article.

Richard Holliday is funded by a National Institute for Health Research Clinical Lectureship. Charlotte Currie is funded by an NIHR Doctoral Research Fellowship. The views expressed are those of the authors and not necessarily those of the NHS, the NIHR or the Department of Health and Social Care.

Thank you to all members of the Child Dental Health Department for their hard work and dedication during restructuring and delivering the service. We are grateful to our colleagues in the Newcastle Dental Hospital Theatre Team and Paediatric Anaesthetics at the Great North Children’s Hospital (Newcastle upon Tyne NHS Hospital Foundation Trust), without whom were could not have delivered a vital component of our service. We would also like to thank Emily Carter for her contribution to the service evaluation.

## Author Contributions

S Simpson contributed to data acquisition and interpretation, drafted, and critically revised the manuscript. C.Currie and R.Holliday, contributed to the conception, design, data acquisition, and interpretation, drafted and critically revised the manuscript. O Sumner, L Burbridge, V Hind, N Lush and B Cole drafted and critically revised the manuscript.

## Notes

### Competing Interest Statement

The authors have declared no competing interest.

### Funding Statement

No funding

### Author Declarations

NUTH Clinical Effectiveness Register reference 1006

